# Towards diagnostic preparedness: detection of highly pathogenic avian influenza A(H5N1) in contrived nasal swab specimens using rapid antigen and point-of-care molecular tests

**DOI:** 10.1101/2025.04.15.25325613

**Authors:** Leda Bassit, Gregory L. Damhorst, Heather B. Bowers, Courtney Sabino, Julie Sullivan, Emily B. Kennedy, Jacob Khouri, Pamela Miller, Eric Lai, Raymond F. Schinazi, Wilbur Lam, Nira R. Pollock, Anuradha Rao

**Affiliations:** Center for ViroScience and Cure, Laboratory of Biochemical Pharmacology, Department of Pediatrics, Emory University School of Medicine, Atlanta, GA, USA; The Atlanta Center for Microsystems-Engineered Point-of-Care Technologies, Atlanta, GA, USA; Division of Infectious Diseases, Department of Medicine, Emory University School of Medicine, Atlanta, GA, USA; OOMVELT, Lakewood, OH 44107, USA; Breton Highlands Consulting, Sand Springs, OK 74063, USA; Echo Consulting, 49 Center Street, Excelsior, MN 55331; PharmaDx, LLC, San Diego, CA 92037, USA; Department of Pediatrics, Emory University School of Medicine, Atlanta, GA, USA; Aflac Cancer and Blood Disorders Center at Children’s Healthcare of Atlanta, Atlanta, GA, USA; Children’s Healthcare of Atlanta, Atlanta, GA, USA; Wallace H. Coulter Department of Biomedical Engineering, Emory University and Georgia Institute of Technology, Atlanta, GA, USA; National Institute of Biomedical Imaging and Bioengineering, National Institutes of Health, Bethesda, MD, USA

## Abstract

**Background:** Highly pathogenic avian influenza (HPAI) A(H5N1) clade 2.3.4.4b was first detected in birds in the United States in 2021 and an ongoing outbreak in dairy cattle began in early 2024. At least 70 U.S. cases have been identified in humans with exposure to infected cattle, poultry, and wild birds. No human-to-human transmission has been documented. However, as part of diagnostic preparedness, we evaluated the ability of currently available influenza tests to detect 2024 U.S. H5N1 strains.

**Methods:** Contrived nasal swab samples were prepared using live or inactivated 2024 H5N1 and used to test twelve rapid antigen tests (lateral flow assays, or LFA), including 10 commercially-available influenza A LFAs and two H5-specific LFAs. Five point-of-care (POC) molecular assays were also tested. An inclusivity testing protocol was used, wherein a predetermined dilution series is used to evaluate each assay, enabling head-to-head comparison of assay performance.

**Results:** All lateral flow assays and POC molecular tests were able to detect bovine 2024 H5N1 (genotype B3.13). Sensitivity for the POC molecular tests (heat-inactivated virus) ranged from 1.55 to 7.75 TCID50/swab. For 11/12 LFAs, including 10 commercial influenza A tests and an RUO H5 assay, sensitivity (live virus) ranged from 78-1550 TCID50/swab. Testing of four LFAs confirmed inclusivity for a genotype D1.1 strain.

**Conclusions:** Available rapid antigen and point-of-care molecular influenza tests can detect 2024 U.S. H5N1 strains in contrived samples, with a wide range of analytical sensitivity. In the event of human-to-human transmission, clinical performance and optimal sample types would need to be established.

**Summary:** Recent human infections with H5N1 influenza necessitate diagnostic preparedness. Reassuringly, commercially available rapid antigen and point-of-care molecular influenza tests detected the 2024 U.S. bovine H5N1 strain in contrived specimens. Clinical performance should be evaluated if human-to-human transmission is observed.

## Introduction

Highly pathogenic avian influenza (HPAI) A(H5N1) has been detected sporadically in humans since 1997 [1]. H5N1 clade 2.3.4.4b, which has been present in birds in the United States since 2021, was identified as the cause of a non-specific syndrome among dairy cattle in the US in early 2024 [2]. Since then, at least 70 confirmed human cases have been identified in the United States, primarily among individuals with exposure to cattle, poultry, or wild birds, including a severe case leading to hospitalization and death [3-5]. Although the public health risk is currently considered to be low, these patterns have prompted preparedness efforts acknowledging the potential for spontaneous mutations or reassortment with seasonal influenza A leading to widespread human-to-human transmission of HPAI H5N1.

The critical role of diagnostics in pandemic preparedness was a major lesson of the SARS-CoV-2 pandemic. The public health response facilitated an acceleration in diagnostic test innovation that shifted testing paradigms from limited centralized testing resources at the onset of the pandemic to widespread point-of-care (POC) and over-the-counter (OTC) diagnostics [6, 7]. This momentum has since also fueled advances in influenza diagnostics, as the first commercial OTC self-tests for influenza A and B have been cleared by the United States Food and Drug Administration (FDA) [8] in the form of multiplexed rapid tests also capable of detecting SARS-CoV-2. Currently, five home use multiplex SARS-CoV-2/influenza antigen assays have FDA Emergency Use Authorization, and three have received marketing authorization through traditional FDA 510k pathways [9-11]. POC molecular tests for influenza have been available since 2015 and generally offer superior sensitivity for the detection of influenza virus infection compared to rapid antigen tests [12, 13].

Diagnostic preparedness for the possibility of widespread H5N1 transmission among humans requires verification that the current strains circulating in cattle and birds are detected by available commercial tests. Many existing OTC/POC assays are expected to detect earlier H5N1 strains on the basis of *in silico* or direct inclusivity testing reported in instructions for use (IFU); however, these data do not report specifically on clade 2.3.4.4b [14]. As an early indicator of the inclusivity of existing OTC and POC tests for the clade currently circulating in U.S. cattle and poultry, and to enable a head-to-head comparison of available tests’ performance for HPAI detection, we tested commercially available assays with contrived specimens prepared with live or inactivated representative HPAI H5N1 strains (A/bovine/Ohio/B24OSU-439/2024 (H5N1), genotype B3.13, and A/Washington/240/2024, genotype D1.1).

## Methods

### Influenza viruses

Tissue culture-propagated HPAI A Virus/bovine/Ohio/B24OSU-439/2024 (H5N1) (GISAID ID-EPI_ISL_19178083) was obtained from Biodefense and Emerging Infections Research Resources Repository (BEI Resources, catalog number NR-59872, lot 70070368). The provided virus was originally isolated from a dairy cow on April 5, 2024, in Ohio, USA and propagated in Madin-Darby canine kidney (MDCK) cells and was provided at a concentration of 3.1E6 TCID_50_/mL determined by cytopathic effect in MDCK cells by the manufacturer. This strain belongs to H5N1 clade 2.3.4.4b and genotype B3.13 and is denoted as “2024 HPAI H5N1” in this manuscript. Another H5N1 clade 2.3.4.4b strain with genotype D1.1(A/Washington/240/2024) (GISAID ID-EPI_ISL_19531299) was obtained from the Centers for Disease Control and Prevention (CDC). This strain was isolated from a sample from a human case in Washington State and is denoted in this manuscript as “2024 HPAI H5N1 D1.1.”

Procedures involving live 2009 H5N1 and 2024 HPAI H5N1 strains, including testing of lateral flow assays (LFAs), were performed in Emory University’s biosafety level (BSL)-3 laboratory using personal protective equipment according to standards set by the CDC and the Emory University Environmental Health and Safety Office. Heat-inactivated 2024 HPAI H5N1 was prepared for testing of POC molecular platforms from the cultured virus stock by heating live virus at 60°C for 30 minutes. Inactivation was confirmed by lack of virus propagation and no evidence of cell death (by crystal violet staining) in MDCK cells infected with 100 µL of ten serial 5-fold dilutions starting at a virus concentration of 3.1E4 TCID_50_/mL. For select LFAs, testing was also performed with an inactivated gamma-irradiated (GIV) version of the BEI strain (NR-59886, lot 70070836). For all strains, the inclusivity protocol described below for LFAs was followed.

Additional testing for select LFAs (Healgen-Rapid Check COVID-19/Flu A&B Antigen Test, ACON Laboratories-Flowflex^®^ Plus COVID-19 and Flu A/B Home Test, and LFA C) was carried out using the following live strains: A/mallard/Wisconsin/2576/2009 (H5N1), A/California/04/2009 (H1N1), A/Ohio/09/2015 (H1N1), A/Tasmania/503/2020 (H3N2), and A/Indiana/08/2011 (H3N2).

### Negative nasal swab matrix preparation

Pooled negative nasal swab matrix (PNSM) was used as diluent for all influenza viruses tested. Details are provided in supplementary methods.

### Contrived specimen preparation

2024 HPAI H5N1 and 2024 HPAI H5N1 D1.1 stocks were thawed in a BSL-3 facility and aliquoted in 50 µL aliquots which were then frozen at -80°C. For LFA testing, aliquots were thawed and diluted to the intended final concentrations in PNSM. Then, using a precision pipette, 50 µL of each dilution was pipetted onto the respective assay swab to simulate collection of a nasal sample, and the assay IFU was followed to test that swab. For molecular assays, 50 µL of heat-inactivated 2024 HPAI H5N1 diluted in PNSM was first pipetted onto the respective assay swab. For the Abbott ID NOW™ assays, the swab with added sample was tested directly, according to the IFU. For assays requiring an intermediate elution of the swab in transport medium, swabs were eluted in 3 mL of UTM® Universal Transport Medium™ (3C047N, Copan Diagnostics). These contrived UTM specimens were aliquoted and frozen to facilitate workflow. UTM specimen aliquots were thawed just before testing. All viral concentrations tested were calculated from the TCID_50_/mL provided by the stock manufacturer.

### LFA inclusivity testing

Inclusivity studies (studies designed to answer the question of whether a test recognizes a different strain of the intended target) were performed according to the routine regulatory procedure by first testing contrived samples with the live virus at 10-fold dilutions [for 2024 HPAI H5N1: 310,000; 31,000; 3,100; and 310 TCID_50_/mL (corresponding to 15,500; 1,550 155; and 15.5 TCID_50_ per swab)], each in triplicate. The lowest dilution for which all three replicates generated a positive result was then used to create three additional two-fold serial dilutions, each of which was tested in triplicate. The lowest dilution detected in all three replicates was defined as the lowest concentration detectable through inclusivity testing. The 10 commercially available Influenza A LFA assays tested in this manner included eight for which the manufacturer granted permission to disclose the device name in the publication and two for which the manufacturer requested that the name be redacted. Two H5-specific LFAs (Healgen H5 RUO and Arbor Vita AV Avantage A/H5N1) were also evaluated using this protocol (see Supplementary Methods for details). All of the LFAs except for one (Quidel Sofia Flu + SARS Antigen FIA) were visually read.

### Molecular assay inclusivity testing

Molecular assays tested included the Cepheid Xpert^®^ Xpress CoV-2/Flu/RSV *plus* test (EUA201505) with Cepheid GeneXpert^®^ systems (a GeneXpert XVI platform, which is equivalent in this context to the GeneXpert Xpress authorized for waived settings, was used). (Note: EUA cartridge reagents and software are identical to the 510k approved product with FDA parent document number K231481). Additional assays evaluated included the Roche cobas® Influenza A/B & RSV (K213822) and Roche cobas® SARS-CoV-2 & Influenza A/B (K223591) with the cobas® Liat® System, and the Abbott ID NOW™ Influenza A&B 2 (K191534) and Abbott ID NOW™ COVID-19 2.0 with Influenza A&B 2 sequential workflow (in which a single patient swab is first tested for SARS-CoV-2 (K221925) and then reflexed to Flu A/B testing; (K191534)). For each assay, PNSM contrived with heat-inactivated 2024 HPAI H5N1 virus at 310, 31, and 3.1 TCID_50_/mL (15.5, 1.55, and 0.155 TCID_50_ per swab) was introduced as a simulated nasal swab and processed according to the assay IFU, with intermediate elution in 3 mL of UTM where indicated.

### CDC assay

The CDC H5 Subtyping assay (IVD kit, K190302) was performed as a research use only (RUO) application according to the IFU. RNA was isolated from 120 µL PNSM spiked with a known quantity of virus using the Qiagen EZ1 DSP Viral Kit (utilizing 280 µL of AVL buffer and 120 µL of elution buffer). For RTqPCR, the Invitrogen SuperScript III Platinum One-Step qRT-PCR System was used, utilizing 5 µL RNA and 20 µL PCR master mix per reaction and an Applied Biosystems 7500 Fast instrument for amplification.

### Digital droplet PCR (ddPCR)

For determination of genomic equivalents (GE)/ml by ddPCR, we used the InfA Primers/Probe from the IVD CDC H5 Subtyping Kit. The One-step RT-ddPCR Advanced Kit for Probes (BioRAD) was used according to the manufacturer’s instructions, with 5 µL of RNA and 17 µL of master mix per reaction.

## Results

Using the inclusivity testing protocol, we tested five commonly used POC molecular tests from three manufacturers, and 10 LFAs (for POC and/or OTC use), all with EUA or 510k clearance, for their ability to detect the 2024 HPAI H5N1 strain as influenza A. As seen in **Figure 1** and **supplementary tables 1 and 2**, all of the POC molecular assays (the Cepheid Xpert^®^ Xpress CoV-2/Flu/RSV *plus* test, the Roche cobas^®^ Influenza A/B & RSV and Roche cobas^®^ SARS-CoV-2 & Influenza A/B for use with the Liat system, and the Abbott ID NOW™ Influenza A &B 2 and Abbott ID NOW™ COVID-19 2.0 with Influenza A & B 2 sequential workflow) detected 2024 HPAI H5N1, with the lowest concentration detected ranging from 1.55 to 7.75 TCID_50_ per swab. Currently, the CDC Influenza Virus Real-time RT-PCR Panel is the only FDA 510k-cleared molecular test for subtyping influenza A(H5) virus and thus provided a useful reference point for POC molecular assay assessment (**Figure 1, supplementary Tables 1 and 2**). Notably, a positive result for H5 on the CDC subtyping assay requires positive results for three targets, including an influenza A target and two H5-specific targets (Supplementary Tables 1 and 2). Data for sample concentrations expressed in genomic equivalents (GE)/mL, as generated by ddPCR, are provided in Supplementary Tables 1 and 2. All 10 LFAs tested detected live 2024 HPAI H5N1, with the lowest concentration detected ranging from 78 TCID_50_ per swab to 1550 TCID_50_ per swab (**Figure 1 and supplementary Table 3**).

**Figure 1:**
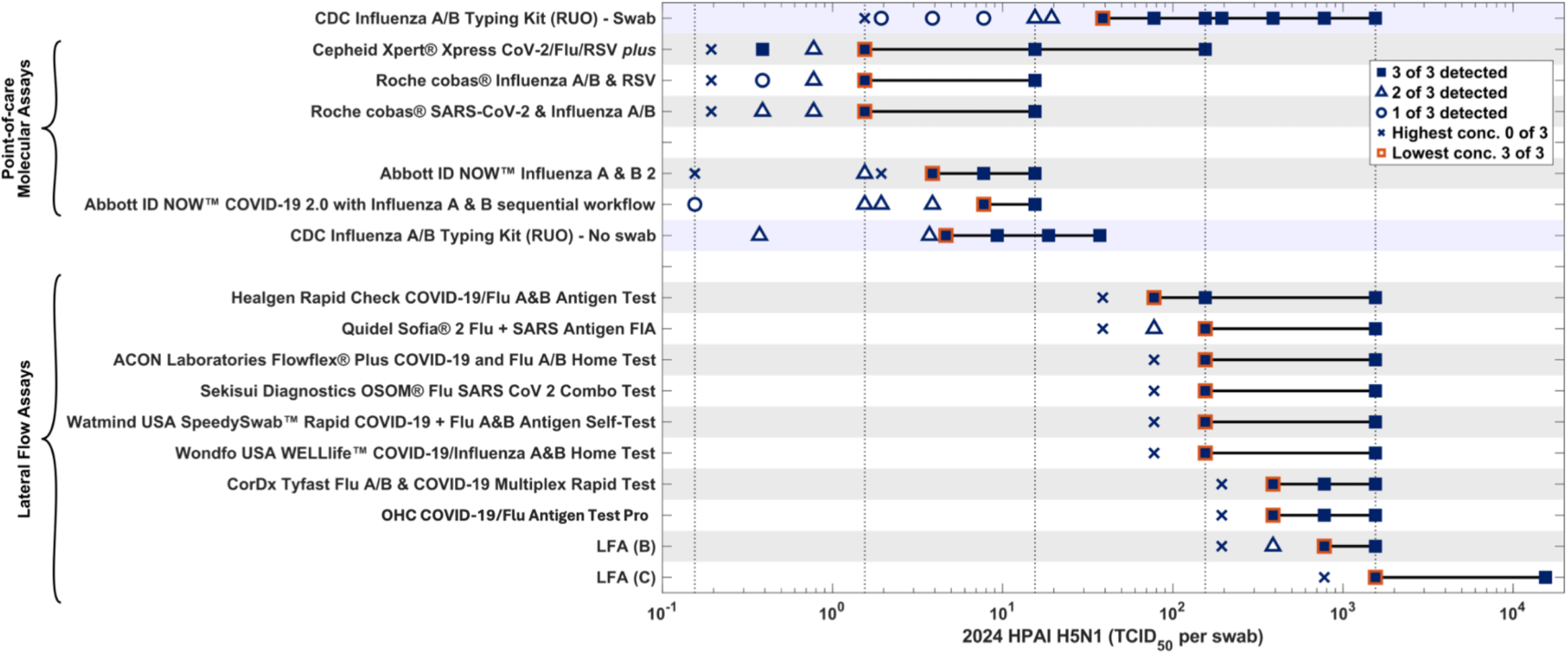
Performance of five POC molecular tests and 10 LFAs for detection of 2024 HPAI H5N1 in contrived nasal swab specimens. Note: The CDC Influenza A/B typing kit (CDC Human Influenza Virus Real-time RT-PCR Diagnostic Panel, Influenza A/B Typing Kit) was performed in two different ways to facilitate direct comparison to POC molecular assays with different formats. For direct comparison to POC molecular assays utilizing swab samples eluted in UTM [Cepheid Xpert^®^ Xpress CoV-2/Flu/RSV *plus* (350 μL) and Roche cobas^®^ Influenza A/B & RSV and Roche cobas^®^ SARS-CoV-2 & Influenza A/B assays (200 μL each)], 120 μL of the same UTM sample was tested with the CDC assay (“Swab”). For direct comparison to the Abbott ID NOW™ Influenza A &B 2 and Abbott ID NOW™ COVID-19 2.0 with Influenza A & B 2 sequential workflow systems (which utilize 50 μL sample in PNSM directly pipetted to a swab), 120 uL of the same PNSM sample was tested with the CDC assay (“No swab”). The first (most concentrated) sample to yield 0 of 3 positive results is indicated by ‘X’. Lower dilutions for which 0 of 3 replicates were positive are indicated in Supplementary Tables 1, 2 and 3. Heat-inactivated virus was used for molecular assays in BSL-1/BSL-2 lab and live virus was used for lateral flow assays in BSL-3 lab. Two LFAs (LFA B and C) were anonymized at the request of the companies. UTM, Universal Transport Media; LFA, Lateral Flow Assay.

Additionally, two LFAs capable of detecting specifically Influenza A(H5) were evaluated. The lowest concentration of 2024 HPAI H5N1 detected by the Healgen – FluA (H5) Ag Rapid Test Cassette (Dropper&Swab) RUO assay was 775 TCID50/swab (15,000 TCID50/mL) (**supplementary table 3**). This H5 RUO test did not show cross-reactivity to either H1N1 or H3N2 strains at the highest possible concentrations (data not shown). The Arbor Vita AV Avantage A/H5N1 Flu Test (utilizing a revised workflow and IFU relative to the 2009 510k version, as provided by the manufacturer) did not detect the most concentrated sample of the 2024 HPAI H5N1 (15,500 TCID50/swab, added to 3 mL M6 transport media). When evaluated with an alternative workflow utilizing viral stock/PNSM mixed 1:1 with M6 (Supplementary Methods), the AV test detected an amount of virus equivalent to 232,500 TCID50 in 3 mL of transport media (77,500 TCID50/mL).

A subset of LFAs (510k and Healgen H5-specific RUO LFA) were tested with gamma-inactivated 2024 HPAI H5N1 available from BEI (Methods), which was generated from the same virus stock used for live virus testing in Figure 1. Results indicated that gamma inactivation does not significantly affect antigenicity of the nucleoprotein (NP) target protein, with the lowest concentration detected by three influenza A LFAs ranging from 39 TCID_50_ per swab to 388 TCID_50_ per swab (**supplementary Table 4**). The hemagglutinin (HA) target (for the H5-specific RUO LFA) appeared to be slightly more affected by gamma irradiation, with the lowest concentration detected increasing 5-fold (from 775 to 3,875 TCID50/swab).

As shown in **Figure 1**, the influenza A LFAs varied in their sensitivity for detection of the 2024 HPAI H5N1 strain. The Healgen Rapid Check COVID-19/Flu A&B Antigen test 510k assay was the most sensitive and detected down to 78 TCID_50_/swab in inclusivity testing, while the least sensitive LFA (LFA C) detected down to 1,550 TCID_50_/swab. **Table 1** compares the relative sensitivity of the LFAs for detection of the 2024 HPAI H5N1 strain using a “relative ratio,” with the sensitivity (lowest concentration detected in inclusivity testing) of the Healgen Rapid Check COVID-19/Flu A&B antigen test (the LFA with De Novo authorization) as the reference (relative ratio = 1). Relative ratios for the sensitivity of detection of 2024 HPAI H5N1 ranged from 2 to 20 (**Table 1**). For comparison, we also evaluated the relative sensitivity of the LFAs for detection of a 2009 H5N1 strain (Mallard/Wisconsin/2576/2009) [Table 1, column H5N1 (2009)]. Inclusivity data for the 2009 H5N1 strain were publicly available for seven LFAs and newly generated at Emory for LFA C. Relative ratios for detection of the 2009 H5N1 strain similarly ranged between 0.2 (Watmind) to 10 (LFA C). The CDC assay again provided a useful reference for determining Ct values in samples used for evaluation of the LFAs (**supplementary Table 3**). **Supplementary Table 3** also indicates ddPCR results (in genomic equivalents/mL) for each sample tested.

**Table 1:**
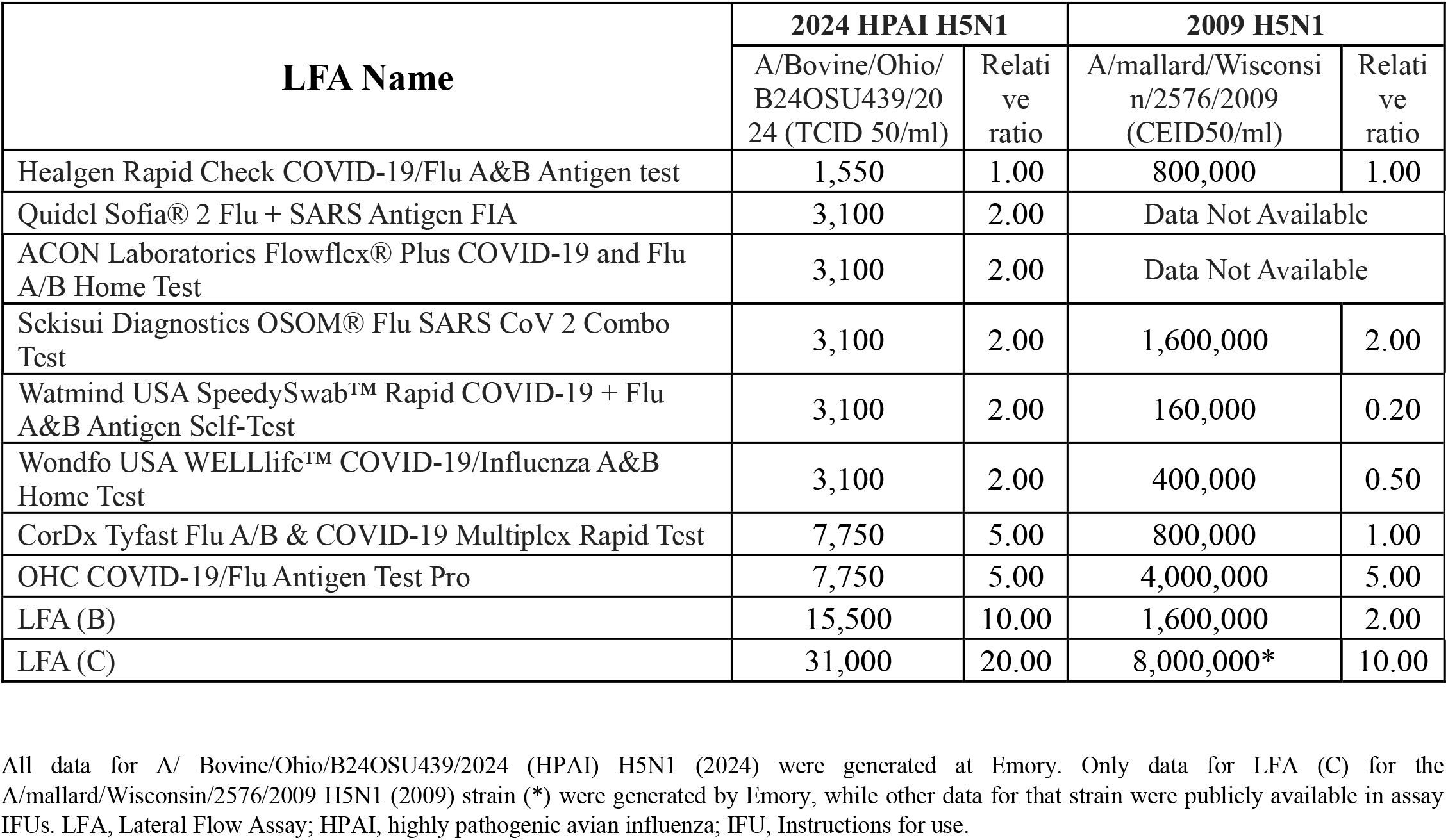
Relative sensitivity of LFAs for detection of 2024 HPAI H5N1 and 2009 H5N1 strains.

| LFA Name | 2024 HPAI H5N1 |  | 2009 H5N1 |  |
| --- | --- | --- | --- | --- |
|  | A/Bovine/Ohio/B24OSU439/2024 (TCID 50/ml) | Relative ratio | A/mallard/Wisconsin/2576/2009 (CEID50/ml) | Relative ratio |
| Healgen Rapid Check COVID-19/Flu A&B Antigen test | 1,550 | 1.00 | 800,000 | 1.00 |
| Quidel Sofia® 2 Flu + SARS Antigen FIA | 3,100 | 2.00 | Data Not Available |  |
| ACON Laboratories Flowflex® Plus COVID-19 and Flu A/B Home Test | 3,100 | 2.00 | Data Not Available |  |
| Sekisui Diagnostics OSOM® Flu SARS CoV 2 Combo Test | 3,100 | 2.00 | 1,600,000 | 2.00 |
| Watmind USA SpeedySwab™ Rapid COVID-19 + Flu A&B Antigen Self-Test | 3,100 | 2.00 | 160,000 | 0.20 |
| Wondfo USA WELLlife™ COVID-19/Influenza A&B Home Test | 3,100 | 2.00 | 400,000 | 0.50 |
| CorDx Tyfast Flu A/B & COVID-19 Multiplex Rapid Test | 7,750 | 5.00 | 800,000 | 1.00 |
| OHC COVID-19/Flu Antigen Test Pro | 7,750 | 5.00 | 4,000,000 | 5.00 |
| LFA (B) | 15,500 | 10.00 | 1,600,000 | 2.00 |
| LFA (C) | 31,000 | 20.00 | 8,000,000* | 10.00 |

All data for A/ Bovine/Ohio/B24OSU439/2024 (HPAI) H5N1 (2024) were generated at Emory. Only data for LFA (C) for the A/mallard/Wisconsin/2576/2009 H5N1 (2009) strain (\*) were generated by Emory, while other data for that strain were publicly available in assay IFUs. LFA, Lateral Flow Assay; HPAI, highly pathogenic avian influenza; IFU, Instructions for use.

To understand whether the observed variations in sensitivity for H5N1 detection were unique to H5N1, we examined the relative sensitivity of the influenza A LFAs for the seasonal flu strains (H1N1 and H3N2) that they were designed to detect (**Table 2**). Importantly, the range and distribution of relative ratios for detection of H1N1 and H3N2 were very similar to the relative ratios for detection of the 2024 HPAI and 2009 H5N1 strains. This indicates that while the LFAs do have some variation in their ability to detect H5N1, they demonstrated very similar and stable variation in their ability to detect the seasonal influenza A strains for which they were designed.

**Table 2:**
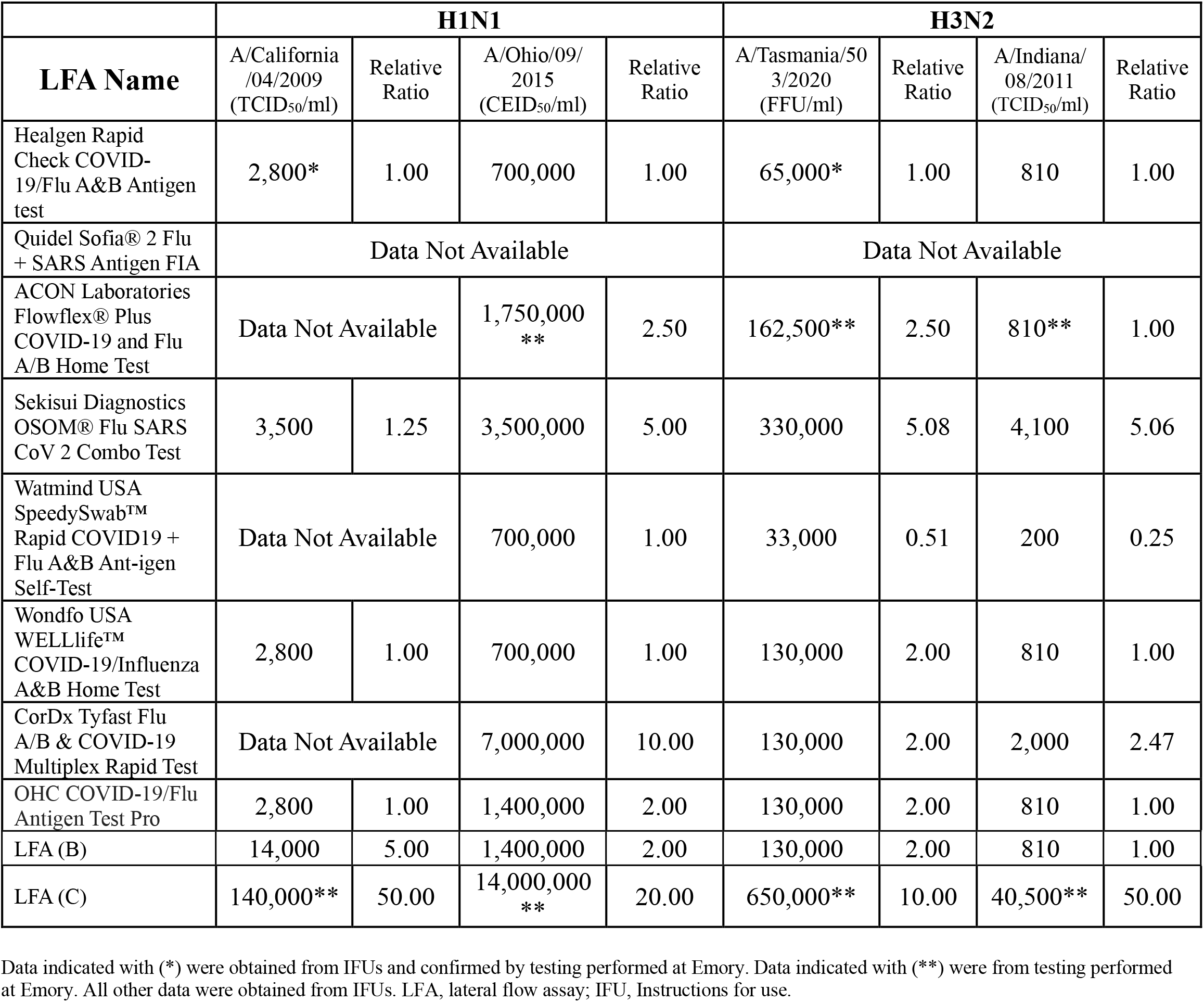
Relative sensitivity of LFAs for detection of seasonal influenza strains (H1N1 and H3N2).

Finally, live 2024 HPAI H5N1 D1.1 (a genotype circulating in poultry and wild birds) was used to test the same four LFAs that were tested with GIV 2024 HPAI H5N1. All four tests were inclusive for 2024 HPAI H5N1 D1.1 (**supplementary table 5**). To estimate relative sensitivity of the Healgen H5-specific RUO LFA for 2024 HPAI H5N1 versus 2024 HPAI H5N1 D1.1, we again calculated relative ratios using the Healgen Rapid Check COVID-19/Flu A&B antigen test as the reference. The relative ratio was 10 for 2024 HPAI H5N1 (**supplementary table 3**), and 0.4 for 2024 HPAI H5N1 D1.1 (**supplementary table 5**), making the Healgen H5-specific RUO assay 25 times more sensitive (10/0.4) for detection of 2024 HPAI H5N1 D1.1.

## Discussion

This study primarily evaluated the ability of commercially available rapid antigen (lateral flow) and molecular influenza A assays for use in POC and/or OTC settings to detect the 2024 HPAI H5N1 strain circulating in dairy cattle in the U.S. All tests selected for the study are “combo” tests, able to detect not only influenza A but also SARS-CoV-2, influenza B, and in some cases, RSV. Tests were compared to the CDC influenza A/H5 assay (implemented as a research application), and the use of consistent reagents and protocols enabled a direct head-to-head comparison of assay performance.

Using the standard inclusivity testing protocol, our data demonstrated that all of the commercially available influenza A LFAs are able to detect the 2024 HPAI H5N1 strain (genotype B3.13) in contrived samples within a 20-fold range of sensitivity. Additional testing of four of the LFAs (510k and Healgen H5-specific RUO LFA) confirmed that they were also able to detect a 2024 HPAI H5N1 D1.1 (a genotype circulating in poultry and wild birds) strain. Notably, the range and relative differences in analytical sensitivity for the 2024 HPAI H5N1 strain were observed to be similar for detection of a 2009 H5N1 strain and seasonal influenza A strains (H1N1 and H3N2), indicating reproducible intrinsic differences in the sensitivity of the tests evaluated. While this range of analytical sensitivity is likely to translate into a range of clinical sensitivity, it is important to note that all the influenza A LFAs evaluated in this study have already been cleared or authorized for the detection of seasonal influenza with support from large prospective clinical trials. All of the POC molecular assays evaluated were shown to be inclusive for the 2024 HPAI H5N1 strain with lowest concentrations detected within a fivefold range. Notably, we also observed that an RUO LFA (Healgen) designed to be specific for H5 detection and optimized for detection of a 2024 H5N1 HA sequence was able to detect both the 2024 HPAI H5N1 and 2024 HPAI D1.1 strains with analytical sensitivity similar to the commercially-available influenza A LFAs, raising the possibility of future use of such a test (once authorized/cleared) either for primary testing or for influenza H5 subtyping following influenza A detection by LFA.

Noting that test manufacturers would benefit from the ability to use inactivated viral stock reagents for test development, we also evaluated the ability of selected tests to detect the BEI gamma-irradiated virus (GIV) derived from the live 2024 HPAI H5N1 strain. As expected, there was no apparent impact of gamma-irradiation on molecular detection with the CDC assay. While the GIV 2024 HPAI H5N1 strain was detected by the influenza A LFAs (which detect a NP target) with sensitivity equivalent to the live strain, we did note that the Healgen H5-specific RUO LFA detected the GIV strain with 5-fold less sensitivity than the live strain, suggesting that developers of tests detecting the HA target should test their assays against live H5 viruses.

These analytical data are key to preparedness, enabling rapid response in the event of increased human-to-human transmission of H5N1. To date, almost all of the US H5N1 cases have been limited to individuals directly exposed to infected animals, and clear evidence of human-to-human transmission has not been observed. The commercial assays evaluated in this study are cleared/authorized for anterior nares (AN) swab specimens or, in the case of the POC molecular assays, AN and nasopharyngeal (NP) swab specimens. The clinical performance of these assays in patients with H5N1 infection, many of whom have presented with conjunctivitis [15, 16], is currently unknown due to a lack of H5N1 clinical samples for evaluation. To date, concentrations of the virus in PCR-positive clinical samples have been relatively low (mean Ct value of 31.0) [16]. However, if there were to be evolution of this virus such that increased human-to-human transmission was observed, the sample types optimal for clinical testing (and viral concentrations in those samples) would need to be defined.

Our inclusivity testing data provide support for our national and global efforts to prepare for H5N1 influenza and demonstrate the utility of expeditious head-to-head evaluation of available tests using consistent reagents and protocols. In the event of an emergence of an evolved strain of H5N1 with higher rates of human-to-human transmission, it would be prudent to evaluate the performance of these assays using the new strain and additional types of clinical samples [17].

## Supporting information

2024 H5N1 Manuscript 04.07.25 Supplemental Material

## Data Availability

All data produced in the present study are available upon reasonable request to the authors

## Funding

The research reported in this publication was supported by the National Institutes of Health contract number [75N92022D00015/75N92023F00001] and grant number [3U54EB027690-05S1] as part of the Rapid Acceleration of Diagnostics (RADx®) initiative.

## Acknowledgments

We wish to thank Joe Brewoo, Kira Moresco, Kalpana Rengarajan for establishing protocols for testing live HPAI in the BSL-3; Nils Schoof, Maud Mavigner for virus inactivation and confirmation of inactivation; Tamara McBrayer, Jerilyn D. Williams for obtaining permits to receive the virus; Mimi Le and the Children’s Clinical and Translational Discovery Core, Department of Pediatrics, Emory University for providing the PNSM used in these studies; and Kaleb McLendon and Joseph Najjar for assistance in testing samples. We also want to thank Adam Samuta, Jose Valdesuso and Michael Wolfson who were instrumental in procuring the virus. We thank the CDC for providing A/Washington/240/2024, and Marie Kirby from CDC for providing the subtyping assays and help with establishing the assay at Emory University CLIA for RUO testing of samples used in the studies described here.

## Author Contributions

**Conceptualization:** LB, GLD, EL, NRP, AR. **Data Curation:** LB, GLD, HB, EL, NRP, AR. **Formal Analysis:** LB, GLD, HB, EBK, JK, EL, NRP, AR **Funding Acquisition:** RFS, WL. **Investigation:** LB, HB, CS, AR. **Methodology:** LB, GLD, PM, EL, NRP. **Project Administration:** JS, EBK, JK, AR. **Supervision Validation:** LB, GLD, EBK, PM, EL, AR. **Writing-Original Draft Preparation:** GLD, NRP, AR. **Writing-Review & Editing:** NRP, GLD, JS, EBK, JK, PM, EL, NP, AR.

## Disclosure of Interest

The authors declare that they have no conflict of interest. Dr. Pollock serves as a scientific advisor to this project on behalf of NIBIB without program management or funding decision responsibilities.

## Notes

### Competing Interest Statement

The authors have declared no competing interest.

### Author Declarations

IRB of Emory University School of Medicine gave ethical approval for this work.

## References

1. Webby RJ, Uyeki TM. An Update on Highly Pathogenic Avian Influenza A(H5N1) Virus, Clade 2.3.4.4b. J Infect Dis 2024; 230(3): 533–42.

2. Burrough ER, Magstadt DR, Petersen B, et al. Highly Pathogenic Avian Influenza A(H5N1) Clade 2.3.4.4b Virus Infection in Domestic Dairy Cattle and Cats, United States, 2024. Emerg Infect Dis 2024; 30(7): 1335–43.

3. Charostad J, Rezaei Zadeh Rukerd M, Mahmoudvand S, et al. A comprehensive review of highly pathogenic avian influenza (HPAI) H5N1: An imminent threat at doorstep. Travel Med Infect Dis 2023; 55: 102638.

4. CDC. H5 Bird Flu: Current Situation. Available at: https://www.cdc.gov/bird-flu/situation-summary/index.html. Accessed 4/07/25.

5. CDC. CDC Confirms First Severe Case of H5N1 Bird Flu in the United States. Available at: https://www.cdc.gov/media/releases/2024/m1218-h5n1-flu.html. Accessed 12/23/24.

6. Santos S, Humbard M, Lambrou AS, et al. The SARS-CoV-2 test scale-up in the USA: an analysis of the number of tests produced and used over time and their modelled impact on the COVID-19 pandemic. The Lancet Public Health 2025; 10(1): e47–e57.

7. Tromberg BJ, Schwetz TA, Perez-Stable EJ, et al. Rapid Scaling Up of Covid-19 Diagnostic Testing in the United States - The NIH RADx Initiative. N Engl J Med 2020; 383(11): 1071–7.

8. FDA. FDA Authorizes Marketing of First Home Flu and COVID-19 Combination Test Outside of Emergency Use Authorities. Available at: https://www.fda.gov/news-events/press-announcements/fda-authorizes-marketing-first-home-flu-and-covid-19-combination-test-outside-emergency-use. Accessed 12/23/2024.

9. USFDA. K243262. Available at: https://www.accessdata.fda.gov/cdrh_docs/reviews/K243262.pdf. Accessed 1/22/2025.

10. USFDA. K243256. Available at: https://www.accessdata.fda.gov/cdrh_docs/reviews/K243256.pdf. Accessed 2/10/2025.

11. USFDA. DEN240029. Available at: https://www.accessdata.fda.gov/cdrh_docs/pdf24/DEN240029.pdf. Accessed 2/26/2025.

12. Babady NE, Dunn JJ, Madej R. CLIA-waived molecular influenza testing in the emergency department and outpatient settings. J Clin Virol 2019; 116: 44–8.

13. Murphy C, Mak L, Cheng SMS, et al. Diagnostic performance of multiplex lateral flow tests in ambulatory patients with acute respiratory illness. Diagn Microbiol Infect Dis 2024; 110(1): 116421.

14. Pinsky BA, Bradley BT. Opportunities and challenges for the U.S. laboratory response to highly pathogenic avian influenza A(H5N1). J Clin Virol 2024; 174: 105723.

15. Uyeki TM, Milton S, Abdul Hamid C, et al. Highly Pathogenic Avian Influenza A(H5N1) Virus Infection in a Dairy Farm Worker. N Engl J Med 2024; 390(21): 2028–9.

16. Garg S, Reinhart K, Couture A, et al. Highly Pathogenic Avian Influenza A(H5N1) Virus Infections in Humans. N Engl J Med 2024.

17. CDC. Conjunctival Swab Specimen Collection for Detection of Avian Influenza A(H5) Viruses. Available at: https://www.cdc.gov/bird-flu/media/pdfs/2024/07/conjunctival-swab-collection-avian-influenza.pdf. Accessed 2/26/2025.

