## Supplementary material for "Towards diagnostic preparedness: detection of highly pathogenic avian influenza A(H5N1) in contrived nasal swab specimens using rapid antigen and point-of-care molecular tests": 2024 H5N1 Manuscript 04.07.25 Supplemental Material

**Supplementary Methods**

*Negative nasal swab matrix preparation*

Pooled negative nasal swab matrix (PNSM) was used as diluent for 2024 HPAI H5N1 and for other influenza viruses tested. Nylon® Flocked Swab (FLOQSwabs® Flocked Swabs 520CS01, Copan Diagnostics) were provided to consented donors (Emory IRB protocol number IRB00089506). Two flocked swabs were used at each time point: first, each swab was used to gently swab one nostril, and then the swabs were crisscrossed to swab the other nostril. One swab (the first) was placed in 3mls UTM® Universal Transport Medium™ (3C047N, Copan Diagnostics) and the other swab (the second) was placed in 1ml saline. After a minimum of 30 minutes, the process was repeated with two more swabs, with each swab being placed in 1ml saline. This process was continued until 16 samples were collected, with at least 30 minutes wait time between collections. The first sample in UTM was tested with the Cepheid Xpert^®^ Xpress CoV-2/Flu/RSV *plus* test. After it was confirmed to be negative for SARS-CoV-2, Flu A, Flu B, and RSV, samples in saline from a minimum of 10 donors were pooled. This pool was designated as PNSM and used for all virus dilutions.

*H5-specific LFA Testing*

The Healgen Influenza A (5) Ag Rapid Test Cassette (Swab) (RUO) is a visually read LFA designed to specifically detect the Influenza A hemagglutinin (HA) H5 target using a nasal swab collected from individuals exhibiting symptoms. The test utilizes monoclonal antibodies raised against recombinant HA generated using sequence from strain A/Texas/37/2024(H5N1). We evaluated this test using the live HPAI H5N1 2024 strain (NR-59872) and by using the manufacturer-provided IFU. Briefly, 50 µL of diluted virus in PNSM was added to the swab and then the swab was added to the extraction tube and rotated six times. After one minute of incubation, the swab was removed, and a dropper was added to the extraction tube so that four drops could be added to the sample well of the cassette. Results were read after a 15-minute incubation.

The AV Avantage™ A/H5N1 Flu Test (Arbor Vita Corporation) is a qualitative rapid immunoassay designed to detect influenza A from throat or nasal swabs collected from individuals exhibiting flu symptoms or from viral cultures for the presumptive laboratory identification of influenza A/H5N1. This test received 510(k) clearance in 2009 for use only in high/medium-complexity laboratories. The test has been modified since the 2009 clearance, with a simpler workflow and an updated IFU. It employs monoclonal antibodies and recombinant proteins containing PDZ domains that recognize, capture, and detect the non-structural protein 1 (NS1). Seasonal FluA and A/H5N1 can be detected and read visually on the same strip. We evaluated this test using live 2024 HPAI H5N1 (NR-59872) first by utilizing the direct swab method, in which 50 µL of diluted virus in PNSM was added to the swab and the swab was put into 3 mL M6 transport media (ThermoFisher) following the clinical specimen protocol recommended by the manufacturer (Manufacturer provided IFU PN1001100, Rev. F). Using a precision pipette, an aliquot of 100µL of the specimen in M6 was transferred to a sample reaction tube and 25µL of sample extraction buffer was then added. After vortexing, 75µL of sample was added directly to the test cassette. Tests were visually read after 45 minutes of incubation using the readout guide provided in the IFU. In addition to the direct swab method, the test was evaluated using the viral culture specimen protocol, following the IFU. In this method, diluted virus in PNSM was added to M6 transport media in a 1:1 volume ratio. A precision pipette was then used to transport 100µL of specimen to a sample reaction tube and the sample was then processed further as described above.

**Supplementary Data**

**Supplementary Table 1: Relative sensitivity of Cepheid Xpert® Xpress CoV-2/Flu/RSV *plus*, Roche cobas® Influenza A/B & RSV nucleic acid test and Roche cobas® SARS-CoV-2 & Influenza A/B nucleic acid test for use with the Liat system for detection of the 2024 HPAI H5N1 strain.**

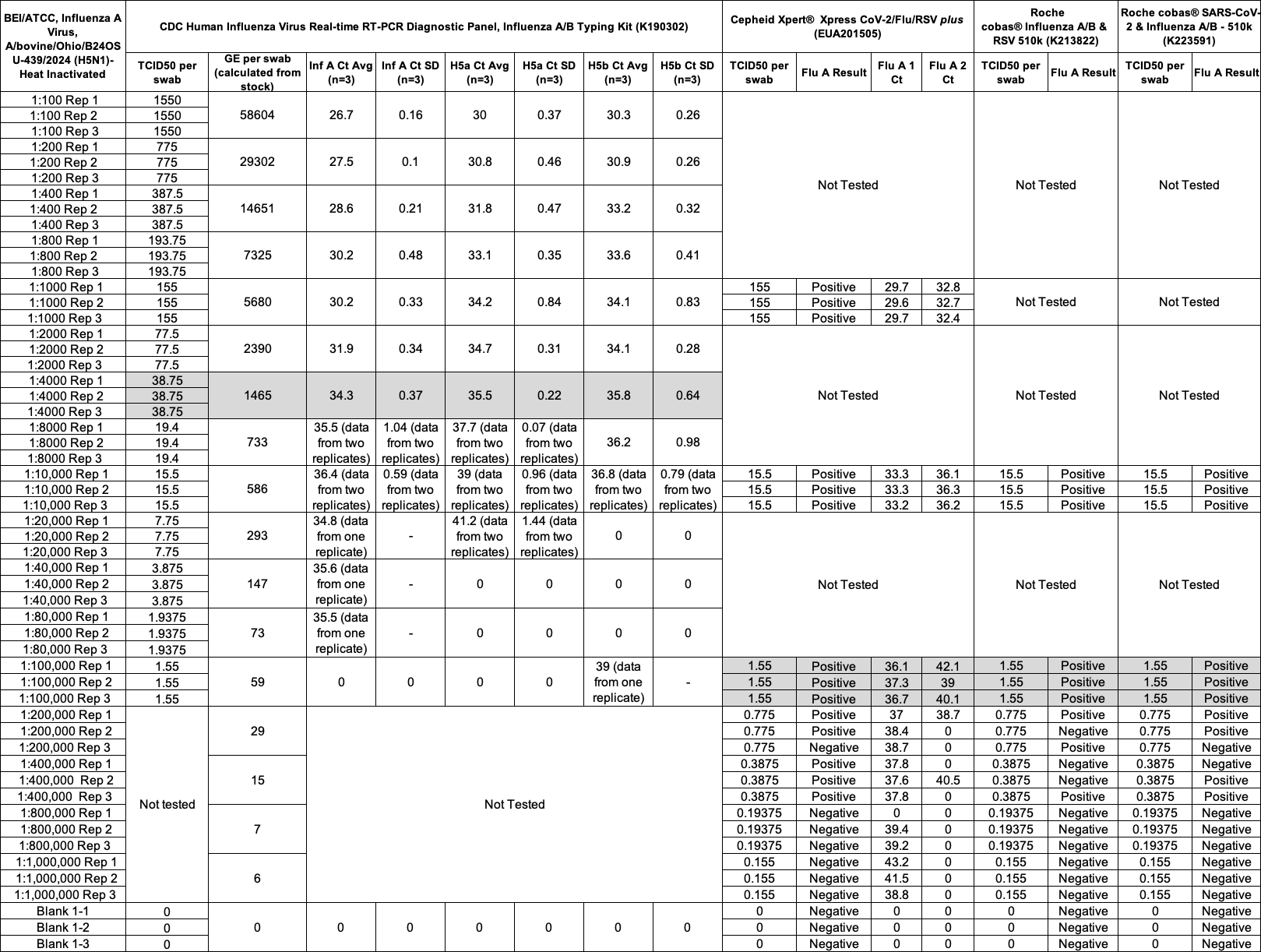

All data were generated using the inclusivity protocol with heat-inactivated 2024 HPAI H5N1. The CDC Human Influenza Virus Real-time RT-PCR Diagnostic Panel, Influenza A/B Typing Kit was performed as a RUO application. For direct comparison of the CDC assay to the Cepheid Xpert^®^ Xpress CoV-2/Flu/RSV *plus* (350 μL) and Roche cobas^®^ Influenza A/B & RSV and Roche cobas^®^ SARS-CoV-2 & Influenza A/B assays (200 μL each), 120 μL of the same UTM sample was tested with the CDC assay. The viral genome equivalent per milliliter (GE/mL) in the stock material as determined at Emory was 1.17E+08 (SD ±9.05E+06) (Avg n=16); GE/mL per swab data were calculated according to the dilution factor. Grey shaded cells indicate the lowest concentration detected in inclusivity testing. RUO, Research Use Only; UTM, Universal Transport Media; GE/mL, Genome Equivalents per milliliter.

**Supplementary Table 2: Relative sensitivity of Abbott ID NOW™ Influenza A&B and Abbott ID NOW™ Influenza A&B Sequential Testing for detection of the 2024 HPAI H5N1 strain.**

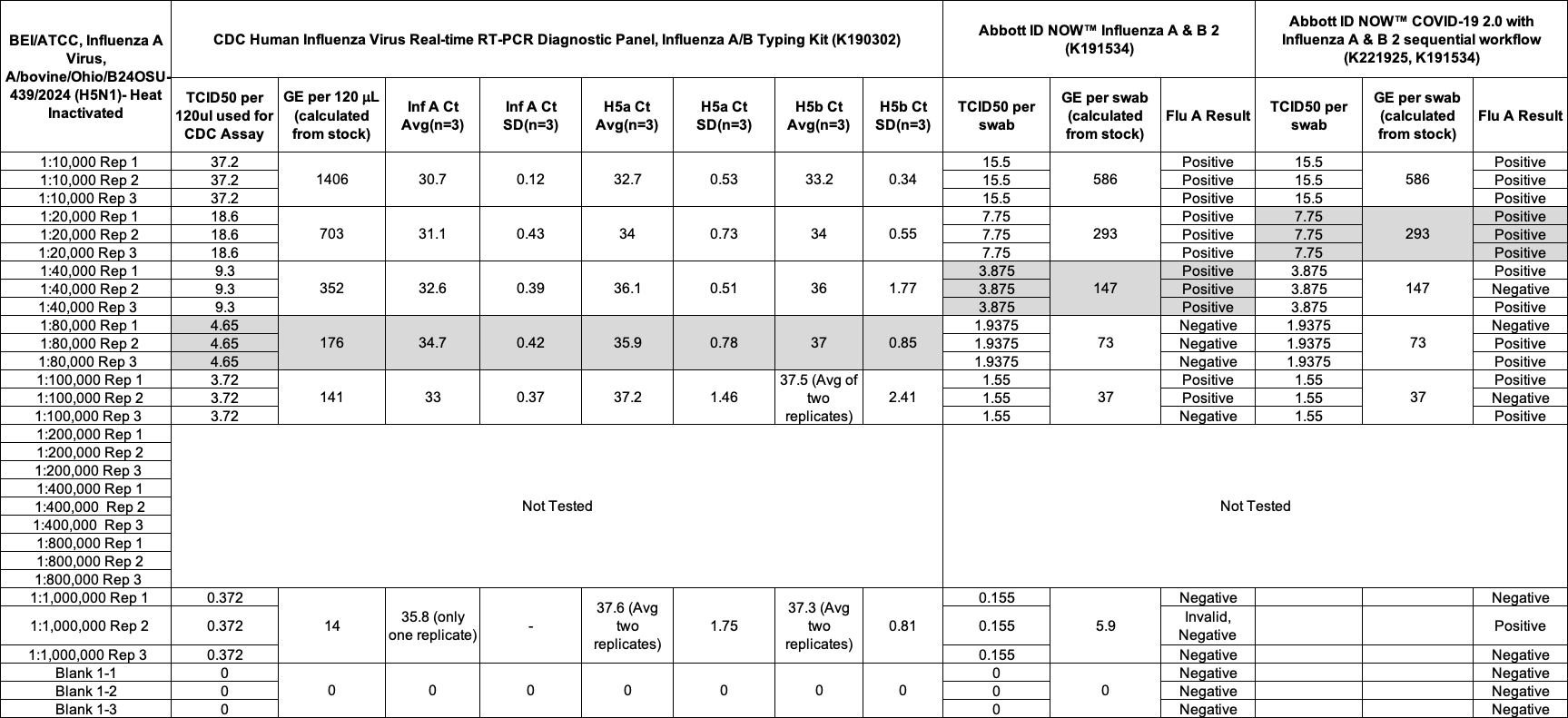

All data were generated using the inclusivity protocol with heat-inactivated 2024 HPAI H5N1. The CDC Human Influenza Virus Real-time RT-PCR Diagnostic Panel, Influenza A/B Typing Kit was performed as a RUO application. For comparison of the CDC assay to the Abbott assays, 120 μL of sample was used for RNA isolation (Methods), as required in the CDC protocol, while for the Abbott assays, 50 μL of sample was pipetted onto a swab, and the swab tested according to the Abbott IFU. The viral genome equivalent per milliliter (GE/mL) in the stock material as determined at Emory was 1.17E+08 (SD ±9.05E+06) (Avg n=16); GE/mL per swab data were calculated according to the dilution factor. Grey shaded cells indicate the lowest concentration detected in inclusivity testing. RUO, Research Use Only; UTM, Universal Transport Media; GE/mL, Genome Equivalents per milliliter.

**Supplementary Table 3: Relative sensitivity of 11 LFAs for detection of the 2024 HPAI H5N1 strain (Live).**

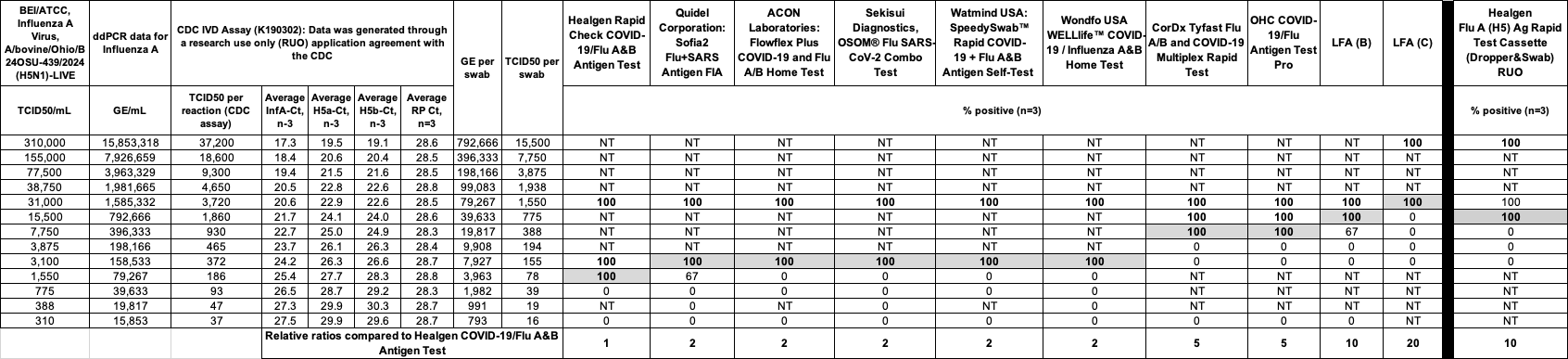

All data were generated using the inclusivity protocol and using live 2024 HPAI H5N1 in the BSL-3. The CDC Human Influenza Virus Real-time RT-PCR Diagnostic Panel, Influenza A/B Typing Kit was performed as a RUO application. For the CDC assay, 120 μL of sample was used for RNA isolation and Ct value determination (Methods). During testing of the LFAs, 50 μL of sample was pipetted onto a swab, and the swab tested according to the assay-specific IFU. The viral genome equivalent per milliliter (GE/mL) for the stock material as determined at Emory is 1.59E+08 (SD ±7.42E+06) (Avg n=16). Grey shaded cells indicate the lowest concentration detected in inclusivity testing. Vertical black column is used to separate data from 10 commercial LFAs and one RUO H5-specific test. For calculation of “relative ratio,” the sensitivity (lowest concentration detected in inclusivity testing) of each test was compared to the sensitivity of the Healgen Rapid Check COVID-19/Flu A&B antigen test (the LFA with De Novo authorization) as the reference (relative ratio = 1). RUO, Research Use Only; NT, Not Tested; InfA, Influenza A; H5a, H5a subtype of influenza A; H5b, H5b subtype of influenza A; Ct, cycle threshold; LFA, Lateral Flow Assay; IFU, Instructions for Use; GE/mL, Genome Equivalents per milliliter.

**Supplementary Table 4: Relative sensitivity of 4 LFAs for detection of the 2024 HPAI H5N1 strain (GIV)**

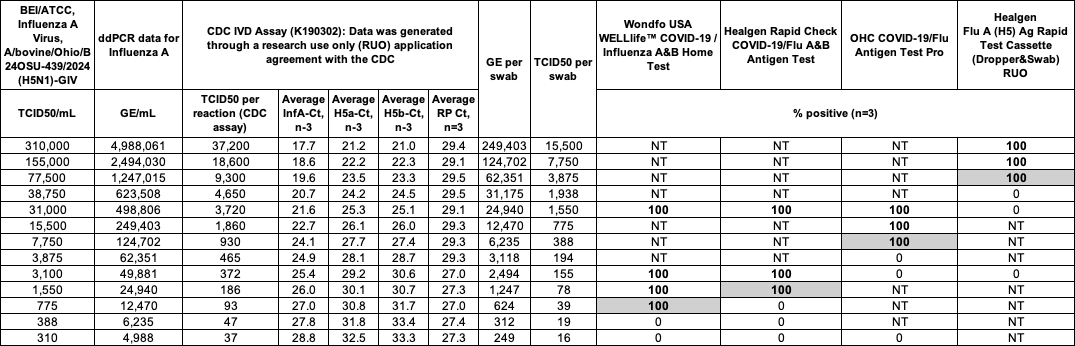

All data were generated using the inclusivity protocol and using GIV 2024 HPAI H5N1. The CDC Human Influenza Virus Real-time RT-PCR Diagnostic Panel, Influenza A/B Typing Kit was performed as a RUO application. For the CDC assay, 120 μL of sample was used for RNA isolation and Ct value determination (Methods). During testing of the LFAs, 50 μL of sample was pipetted onto a swab, and the swab tested according to the assay-specific IFU. The viral genome equivalent per milliliter (GE/mL) of the stock material as determined at Emory is 4.99E+07 (SD ±1.19E+06) (Avg n=16). Grey shaded cells indicate the lowest concentration detected in inclusivity testing. GIV, Gamma Inactivated Virus; RUO, Research Use Only; NT, Not Tested; InfA, Influenza A; H5a, H5a subtype of influenza A; H5b, H5b subtype of influenza A; Ct, cycle threshold; LFA, Lateral Flow Assay; IFU, Instructions for Use; GE/mL, Genome Equivalents per milliliter.

**Supplementary Table 5: Inclusivity testing of 4 LFAs for detection of the 2024 HPAI H5N1 D1.1 strain**

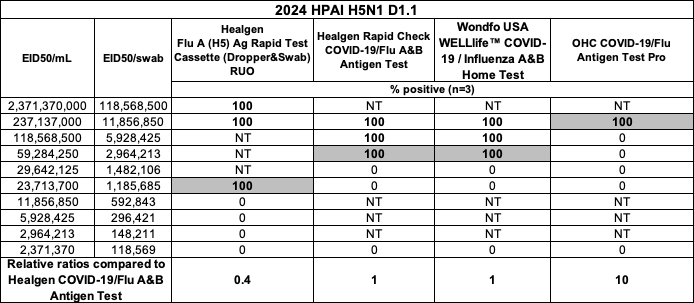

All data were generated using the inclusivity protocol and 2024 HPAI H5N1 D1.1. During testing of the LFAs, 50 μL of sample was pipetted onto a swab, and the swab tested according to the assay-specific IFU. Grey shaded cells indicate the lowest concentration detected in inclusivity testing. For calculation of “relative ratio,” the sensitivity (lowest concentration detected in inclusivity testing) of each test was compared to the sensitivity of the Healgen Rapid Check COVID-19/Flu A&B antigen test (the LFA with De Novo authorization) as the reference (relative ratio = 1). LFA, Lateral Flow Assay; RUO, Research Use Only; EID, Egg Infectious Dose; NT, Not Tested.
